# *Galar* - a large multi-label video capsule endoscopy dataset

**DOI:** 10.1101/2024.09.26.24314265

**Authors:** Maxime Le Floch, Fabian Wolf, Lucian McIntyre, Christoph Weinert, Albrecht Palm, Konrad Volk, Paul Herzog, Sophie Helene Kirk, Jonas L. Steinhäuser, Catrein Stopp, Mark Enrik Geissler, Moritz Herzog, Stefan Sulk, Jakob Nikolas Kather, Alexander Meining, Alexander Hann, Steffen Palm, Jochen Hampe, Nora Herzog, Franz Brinkmann

## Abstract

Video capsule endoscopy (VCE) is an important technology with many advantages (non-invasive, representation of small bowel), but faces many limitations as well (time-consuming analysis, short battery lifetime, and poor image quality). Artificial intelligence (AI) holds potential to address every one of these challenges, however the progression of machine learning methods is limited by the avaibility of extensive data. We propose *Galar*, the most comprehensive dataset of VCE to date. *Galar* consists of 80 videos, culminating in 3,513,539 annotated frames covering functional, anatomical, and pathological aspects and introducing a selection of 29 distinct labels. The multisystem and multicenter VCE data from two centers in Saxony (Germany), was annotated framewise and cross-validated by five annotators. The vast scope of annotation and size of *Galar* make the dataset a valuable resource for the use of AI models in VCE, thereby facilitating research in diagnostic methods, patient care workflow, and the development of predictive analytics in the field.

## Background & Summary

Video capsule endoscopy (VCE) is a minimally invasive gastroenterological imaging procedure used to capture video footage of a patient’s digestive tract. This is especially relevant for the small intestine, which is not readily accessible through conventional endoscopic procedures like colonoscopy and esophagogastroduodenoscopy. However, this comes with limitations such as a time-consuming manual analysis^1^, technical restrictions (e.g., battery runtime^2^ or a lack of active locomotion), and heterogeneous image quality. In 16.5% of cases, the capsule does not pass through the ileocecal valve, resulting in incomplete small intestine examinations^3^.

VCE is currently primarily employed for the detection of internal bleeding^4,5^. However, the potential use cases for VCE are far more expansive. The indications for capsule endoscopy are evolving alongside technological advancements, such as the introduction of colon capsule endoscopy^6^, thereby expanding its use e.g. in pediatric populations and for inflammatory bowel disease^7^.

The major drawback of VCE is the large amount of video footage generated, as medical staff are required to watch hours of recorded video. In these recordings, the section of interest is a tiny subset of the total video, and fluctuating image quality renders large parts unusable for diagnostic purposes. The use of Artificial intelligence (AI) in VCE is already reducing the diagnostic evaluation time needed to interpret the large amount of VCE footage. With the rise of AI in VCE, the procedure has the potential to become more widely used and thereby more cost-efficient, as observed in other modalities, such as AI-assisted X-Ray evaluation^8^. Lately, the integration of Edge AI emphasizes the growing need for efficient, miniaturized algorithms for low-power devices^9,10^, which opens up new possibilities for real-time analysis within VCE systems.

The successful development of AI models requires substantial quantities of high-quality data, as well as precise and rigorous annotations^11^. However, the availability of large datasets is scarce; the VCE-datasets thus far publicly available are either relatively small^12,13,14^ or are limited to specific questions (e.g., quality, ulcers, bleeding, polyps, anatomy)^12,15,16,17,18^. To further drive the progress of AI in VCE, the creation of large, preprocessed, and annotated datasets is necessary^16,19^. Most academic research projects process their own data, which is tailored to their specific tasks^20,21^ and do not make their datasets publicly available. The drawback of such an individualistic approach is that it necessitates a disproportionate amount of resources, limiting the progress of research. The existence of large, high-quality datasets could reduce the cost and effort involved in developing research for VCE and other medical technologies^22^.

In this publication, we introduce a dataset that marks a considerable advancement in the field of capsule endoscopic research. In the domain of VCE there are a few openly accessible datasets, an overview of these is given in Table 6. Galar positions itself to be one of the largest datasets in the field. With 29 distinct labels, incorporating a broad range of functional, anatomical, and pathological annotations across 3,513,539 frames, Galar is primed for application in the field of machine learning.

Furthermore, the *Galar* dataset consists of VCE data from two endoscopy centers in Germany, with two different capsule systems (Olympus™ Endocapsule 10 System, PillCam™ SB2, SB3, and Colon2 Capsule Endoscopy Systems^23,24^). As multidisciplinary and multicenter VCE research is needed for the clinical use of AI in patient diagnosis^16,25^, this further elevates Galar in its usefulness.

In summary, we provide a multicentric, multisystem dataset with high frame count and the most diverse and detailed annotations to date. These characteristics establish *Galar* as a robust resource for training machine learning models in video capsule endoscopy.

## Methods

Videos were collected from the University Hospital Carl Gustav Carus (Dresden, Germany) and from an outpatient practice for gastroenterology (Dippoldiswalde, Germany). VCE recordings were obtained from August 2011 to March 2023 using the Olympus™ Endocapsule 10 System (Hamburg, Germany) as well as the PillCam™ SB2, SB3, and Colon Capsule Endoscopy Systems (Meerbusch, Germany)^23,24^. The videos were initially generated in proprietary data formats and were converted to the Moving Picture Experts Group (MPEG) format. The video resolution ranged from 336 x 336 pixels (Olympus™) to 576 x 576 pixels (PillCam™). Out of the 449 recordings, 80 videos were pre-selected for annotation based on the related findings by selecting only pathological videos for annotation. To de-identify VCE recordings, randomly generated study IDs were assigned, and the videos were cut. Afterwards, videos were transferred to university servers. There each video in the dataset was labeled framewise, resulting in 3,513,539 labeled frames.

This study was approved by the Ethics Committee of the University Hospital Carl Gustav Carus at the Technical University of Dresden on December 16, 2022 (Ethics ID: BO-EK-534122022), confirming adherence to the ethical principles of the Declaration of Helsinki. Due to the retrospective anonymization of the data and their collection during clinically indicated routine interventions, explicit consent was not required. This is additionally supported by the Ethics Committee’s approval, a consultation with the data privacy officer, and local law. Section 34, Paragraph 1 of the Saxon Hospital Act (SächsKHG) explicitly allows the collection and analysis of this type of data.

### Data preparation

Computer vision annotation tool (CVAT)^26^ is a web-based, open-source image- and video annotation tool. Using CVAT, five annotators (a team of experienced gastroenterologists and trained medical students) labelled the data. The labels were categorized into three main groups: The **technical** group consists of labels concerning the image quality, where *good view* indicates a reduction of the view by less than 50%, *reduced view* indicates a reduction of the view by over 50%, and *no view* indicates a reduction of the view by over 95%. Furthermore, a distinction is made between *bubbles* and *dirt* as factors contributing to the degradation of image quality. The **anatomical** group consists of typical landmarks: *z-line, pylorus, papilla of Vater, ileocecal valve* and the different sections of the gastrointestinal tract: *mouth, esophagus, stomach, small intestine, colon*. The final group is the **pathological** group, which consists of the most frequent pathologies found in VCE and some less frequent findings: *ulcer, polyp, active bleeding, blood, erythema, erosion, angiectasia, inflammatory bowel disease (IBD), foreign body, esophagitis, varices, hematin, celiac, cancer, lymphangiectasis*. The pathologies *esophagitis, varices and celiac* did not occur in any of the videos. Figure 1 gives an overview of example images of the 26 labels in the dataset, Figure 2 displays the number of annotated frames per label.

**Figure 1.**
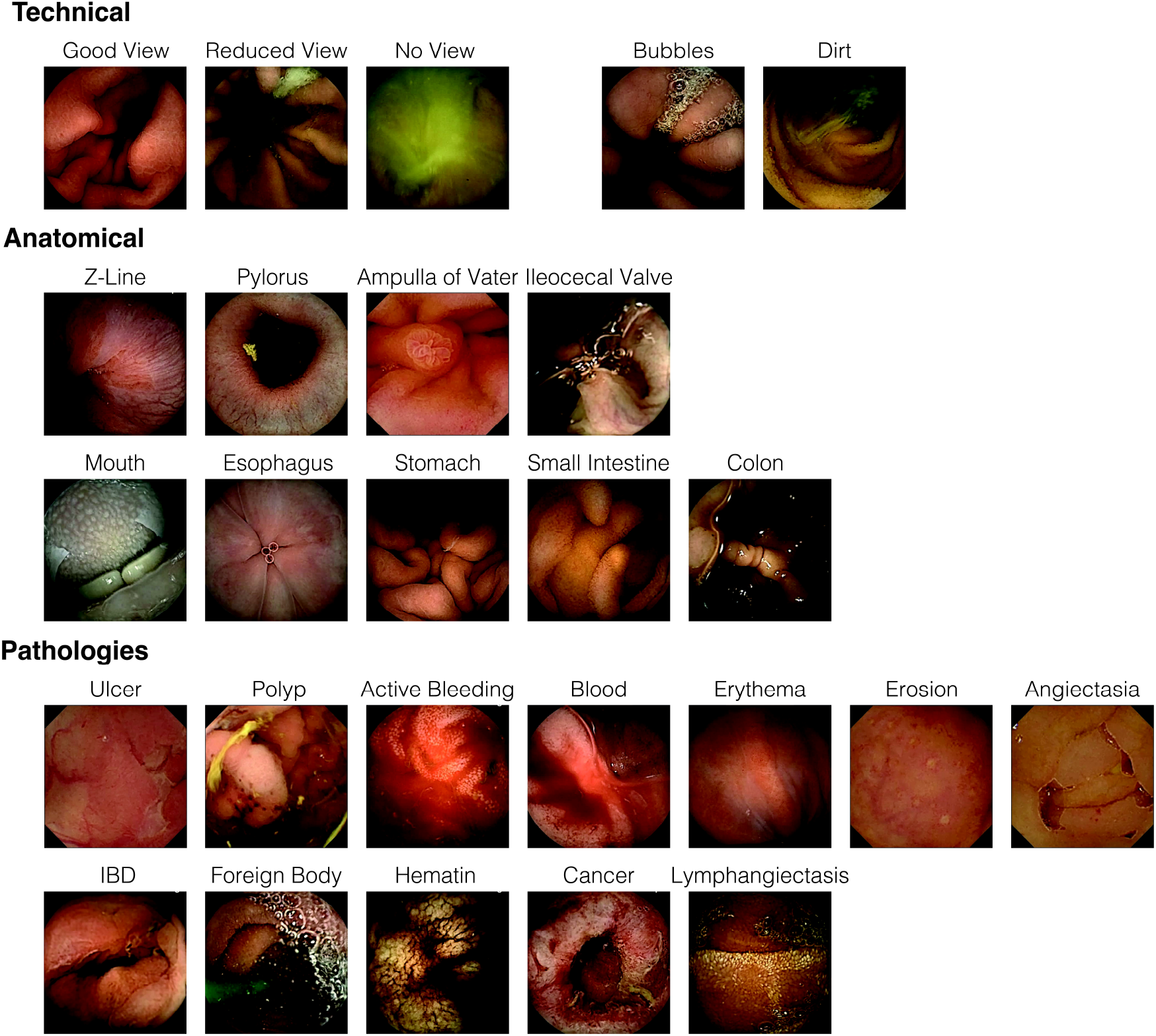
Example images of the 26 labels in the dataset. The figure does not contain images of the labels *esophagitis, varices* and *celiac*, as there were no instances of these pathologies present in the set of VCE studies.

**Figure 2.**
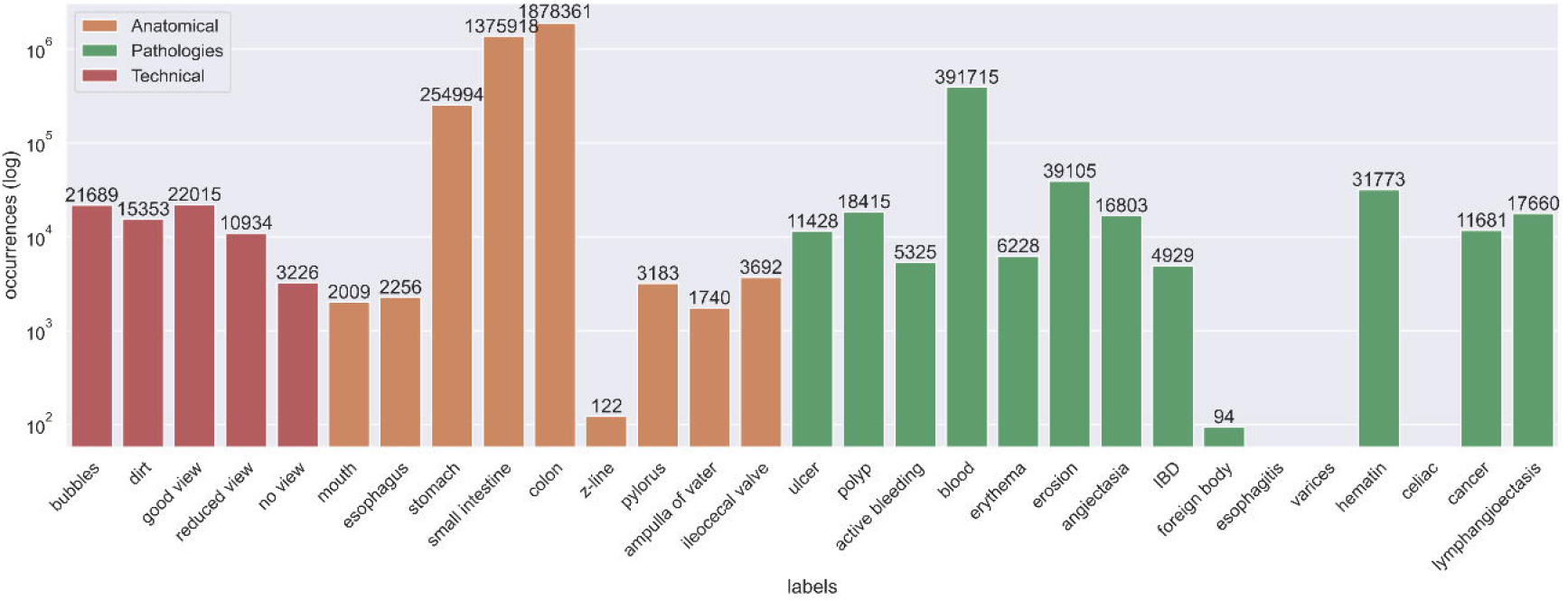
Overall frames per label count of the *Galar* dataset. Image occurrences per labels are displayed across the three main groups (technical, sections and anatomical). The y-axis is scaled logarithmically. Legend: Orange – Anatomical Green – Pathologies Red - Technical

### Annotation Process

An early decision was made to label each frame in the dataset individually, with the annotation process occurring in multiple stages. From each of the videos, every unique frame was extracted using Python (v3.9.8)^27^ and FFMPEG (v4.0.6)^28^. Frames originating from the PillCam™ capsule system were cropped to remove black borders. A timestamp, visible in the top right corner, was also removed. No further pre-processing was done for the videos from the Olympus™ capsule system. Subsequently, the frames were uploaded to CVAT, where frames were annotated by our team. Frames containing unrecognizable features were given the label *unknown*. Then, all frames labeled with a pathology were cross-validated with the confirmation of a secondary annotator. Any frames still possessing the *unknown* label were reviewed by a gastroenterologist with 10 years of experience in endoscopy and were relabeled accordingly.

## Data Records

The *Galar* VCE dataset can be found in the open access repository *figshare*^29^. It consists of 3,513,539 frames, each labeled with 29 labels, and has a total size of ∼580GB. A detailed overview of the structure of the dataset is shown in Figure 3. Each video in the dataset was labeled framewise. The dataset contains the folders *Frames* and *Labels*. The *Labels* folder contains CSV files, where each file has a header starting with the *index* column, followed by the columns of the 29 possible labels described in Data preparation and ending with the *frame* column, which refers to the corresponding frame the labels belong to. The *Frames* folder consists of 80 sub-folders, each containing the frames associated with a study. Table 1 shows the number of videos, resolution, and distribution of frames per capsule system. Additionally, a metadata file is provided, containing patient age, gender, and capsule system used.

**Table 1.** Overview of the data records in the *Galar* dataset. Description of the resolution, number of frames, and number of videos per capsule system.

| Capsule System | Resolution (Pixels) | No. of frames | No. of videos |
| --- | --- | --- | --- |
| PillCam<sup>TM</sup> University Hospital Dresden | 512x512 | 528,470 | 38 |
| Olympus<sup>TM</sup> University Hospital Dresden | 336x336 | 2,750,514 | 22 |
| PillCam<sup>TM</sup> Dippoldiswalde | 512x512 | 234,555 | 20 |

**Figure 3.**
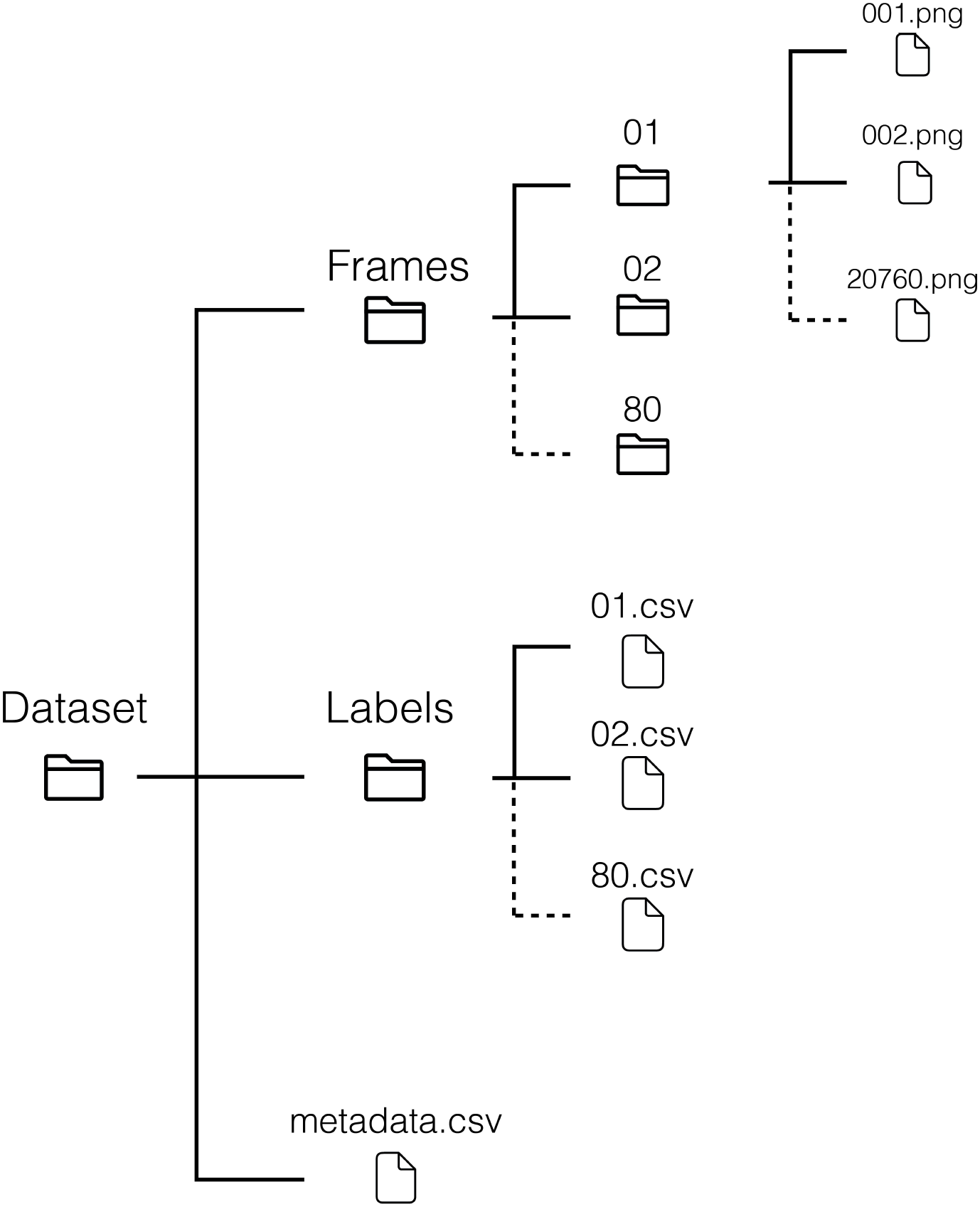
The file structure of the *Galar* dataset. Frames are stored chronologically in subfolders of the *Frames* folder. Labels are stored in a single CSV file, per study. The metadata file further contains data on a per study basis.

Six videos contain technical annotations: *5, 8, 9, 13, 14, 22*. A total of 35,733 frames were annotated with this label category. The creation of technical annotations was found to be more resource intensive compared to the other categories, as the visibility is more volatile and prone to sudden change. As the category is highly relevant for machine learning (ML) applications in VCE, the labels were included for completeness.

## Technical Validation

The dataset was used to train multiple ResNet-50 models^30^. The data was split into a set for training and validation consisting of 60 videos, and a test set comprised of the remaining 20 videos. K-fold cross-validation was performed on the data in the training and validation set. The videos from the test set all originate from the Dippoldiswalde practice and there is no overlap of these videos with those from the train set.

The labels *dirt* and *bubbles* form a multi-label classification problem, while the labels *good view, reduced view*, and *no view* as well as the section labels *mouth, esophagus, stomach, small intestine*, and *colon* require multi-class classification. Some of the other more frequently occurring pathological (e.g., *blood* or *polyp*) labels are trained on separately.

For the classification of the multi-label and the multi-class models, 5-fold cross-validation was employed. The binary classification of the pathologic labels was done using 2-fold cross-validation, as some labels were not contained in a sufficient number of videos. To ensure that the frames of one patient are not spread over the training and test set and to get the best possible distribution of the labels over the folds, sklearn’s StratifiedGroupKFold^31^ method was applied.

The ResNet-50 model pre-trained on ImageNet^32^ was fine-tuned for 10 epochs using PyTorch (v2.0.1)^33^. Following this, fine-tuning was done for each of the target tasks. These models were trained over 100 epochs (with early stopping), with a 128 Batch size and a 0.001 learning rate. For each image, a Resize transform was applied, to scale the image down to 224×224. Additionally, the transforms ShiftScaleRotate, RGBShift, GaussNoise and RandomBrightnessContrast were each applied with a 30% likelihood to each image. The small subset of images which contain the technical annotation were fine-tuned similarly, excepting the epochs, which were capped at 50 with early stopping.

As measurements for the classification performance, the F-1 score, the Area under the Receiver Operating Characteristic Curve (AUROC) as well as the accuracy were calculated using the TorchMetrics (v1.0.3)^34^ Python library.

Tables 2, 3, 4, and 5 show results for the classification models. The model fine-tuned for *dirt* and *bubbles*, along with the two multi-class models, performed decently with accuracy value up to 93% for the labels *stomach* and 92% for *small intestine*.

**Table 2.** Classification results for a ResNet-50 fine-tuned on *bubbles* and *dirt*. The metrics were computed individually for each label, and both macro- and micro-averaged scores are calculated across all labels. The outcomes are averaged across the 5 cross-validation folds.

| Label | F1 | AUROC | Accuracy |
| --- | --- | --- | --- |
| bubbles | 0.89 | 0.87 | 0.82 |
| dirt | 0.88 | 0.94 | 0.88 |
| macro average | 0.88 | 0.90 | 0.85 |
| micro average | 0.89 | - | 0.85 |

**Table 3.** Classification results for a ResNet-50 fine-tuned on *good view, reduced view*, and *no view*. The metrics are computed individually for each label, and both macro- and micro-averaged scores are calculated across all labels. The outcomes are averaged across the 5 cross-validation folds.

| Label | F1 | AUROC | Accuracy |
| --- | --- | --- | --- |
| good view | 0.88 | 0.88 | 0.87 |
| reduced view | 0.41 | 0.85 | 0.47 |
| no view | 0.33 | 0.91 | 0.29 |
| macro average | 0.54 | 0.88 | 0.54 |
| micro average | 0.79 | - | 0.79 |

**Table 4.** Classification results for a ResNet-50 fine-tuned on *mouth, esophagus, stomach, small intestine*, and *colon*. The metrics are computed individually for each label, and both macro- and micro-averaged scores are calculated across all labels. The outcomes are averaged across the 5 cross-validation folds.

| Label | F1 | AUROC | Accuracy |
| --- | --- | --- | --- |
| mouth | 0.42 | 1.00 | 0.75 |
| esophagus | 0.65 | 1.00 | 0.73 |
| stomach | 0.78 | 0.93 | 0.93 |
| small intestine | 0.93 | 0.95 | 0.92 |
| colon | 0.75 | 0.96 | 0.72 |
| macro average | 0.71 | 0.96 | 0.81 |
| micro average | 0.89 | - | 0.89 |

**Table 5.** Classification results for multiple ResNet-50 models, each fine-tuned, on points of interest (e.g. *blood)*. The metrics are computed individually for each label. The outcomes are averaged across the 2 cross-validation folds.

| Label | F1 | AUROC | Accuracy |
| --- | --- | --- | --- |
| blood | 0.14 | 0.87 | 0.98 |
| pylorus | 0.01 | 0.74 | 0.99 |
| z-line | 0.02 | 0.87 | 1.00 |
| ulcer | 0.00 | 0.44 | 0.99 |
| polyp | 0.05 | 0.73 | 1.00 |
| erythema | 0.02 | 0.70 | 1.00 |

**Table 6.**
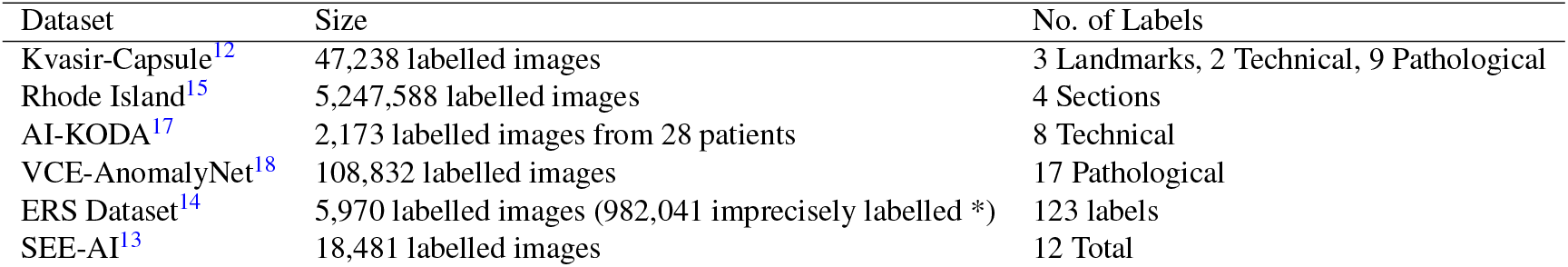
Overview of openly accessible VCE datasets. *Imprecisely labelled images inherit labels from those that are labelled by an expert, and where the image appears chronologically close.

The binary models for pathological labels encountered challenges to accurately identify positive samples. To improve performance, weighted sampling as well as weighted loss was explored. For weighted sampling, the probability for an image to be sampled was based on the occurrence of its class, as a fraction of the total dataset. For the more complex multi-label problem, each unique combination of labels was assigned a weight, again as a portion of the total dataset. This made the weights dynamic, based on the target the model is trained on.

Although these strategies helped to improve performance on some labels, other required heavy parameter optimization. This underscores the difficulty and necessity of improving and developing AI methods to address the challenges of imbalanced label distrubution and multi-source data. Consequently, it highlights the importance of a multicentric, multisystem dataset with extensive annotations of pathologies.

## Usage Notes

With *Galar* we provide the largest public VCE dataset, both in terms of the number of features labeled per image and the total number of annotated images. The large number of ground truth labeled images allows for supervised training of machine learning models and is a significant contribution to the landscape of publicly available VCE datasets.

If the dataset is to be employed for machine learning applications, it is essential to carefully partition the data into training and validation sets. The comparative rarity of select labels, especially over others in the same class, must be respected. Additionally, the data originates from two different VCE systems and was collected at two different study sites. Patients of varying age and gender are also present in the dataset. This information must be considered when generating splits. The metadata file, found in the figshare repository, provides information regarding the capsule system and patient age and gender, per individual study.

The dataset is provided compressed, in the 7-Zip (.7z) format. The data must be uncompressed before it may be viewed and modified; common operating systems (Windows, Linux, MacOs) by default provide archive utility which enables this.

By licensing the dataset under a Creative Commons Attribution 4.0 International (CC BY 4.0) License which allows sharing, copying, and redistribution, as well as adaptation and transformation, we hope to advance research in the field. For more details about Creative Commons licensing, please refer to https://creativecommons.org.

## Data Availability

All data produced in the present study are available upon reasonable request to the authors. Data will be published and openly available upon publishing of the manuscript

## Code availability

The code employed for the technical validation can be accessed via our public GitHub Repository: https://github.com/EKFZ-AI-Endoscopy/GalarCapsuleML. The repository contains a full guide on running the code, tuning hyperparameters, and generating statistics.

## Author contributions statement

M.L.F.: methodology, investigation, data curation, writing original draft, visualization. F.W.: methodology, software, technical validation, writing original draft, visualization. L.M.: methodology, software, data curation, writing original draft, visualization. C.W.: methodology, data annotation. A.P.: methodology, data annotation. K.V.: methodology, data annotation. P.H.: data curation, investigation. S.H.K.: methodology, investigation, clinical supervision. J.L.S.: methodology, investigation, review and editing. C.S: data curation. M.E.G: methodology, data curation, data annotation. M.H.: review and editing. S.S: data curation, technical and clinical supervision. A.M.: methodology, review and editing. A.H.: methodology, review and editing. J.N.K: review and editing, supervision. S.P: conceptualization, investigation, data curation. J.H: conceptualization, review and editing, supervision, N.H: conceptualization, investigation, data curation, writing original draft, review, and editing, visualization, supervision, project administration, funding acquisition. F.B.: conceptualization, investigation, data curation, review, and editing, visualization, supervision, project administration, funding acquisition.

This research was funded by the BMBF (German Federal Ministry of Education and Research) as part of the SEMECO cluster4future FKZ 03ZU1210GA and 03ZU121HB.

